# Monosomy 7/del(7q) Cause Sensitivity to Inhibitors of Nicotinamide Phosphoribosyltransferase in Acute Myeloid Leukemia

**DOI:** 10.1101/2023.04.08.23287765

**Authors:** Samuli Eldfors, Joseph Saad, Nemo Ikonen, Disha Malani, Bjørn Tore Gjertsen, Mika Kontro, Kimmo Porkka, Caroline A. Heckman

**Affiliations:** Institute for Molecular Medicine Finland, Helsinki Institute of Life Science, iCAN Digital Precision Cancer Medicine Flagship, University of Helsinki, Helsinki, Finland; Department of Internal Medicine, Faculty of Medicine, University of Helsinki, Helsinki, Finland; Research Program in Systems Oncology, Faculty of Medicine, University of Helsinki, Helsinki, Finland; Massachusetts General Hospital Cancer Center, Charlestown, MA, USA; Harvard Medical School, Boston, MA, USA; Department of Medical Oncology, Dana-Farber Cancer Institute, Boston, MA, USA; Department of Medicine, Hematology Section, Haukeland University Hospital, Bergen Norway; Center for Cancer Biomarkers, Department of Clinical Science, University of Bergen, Bergen, Norway; Department of Hematology, Helsinki University Hospital Comprehensive Cancer Center, Helsinki, Finland; Foundation for the Finnish Cancer Institute, Helsinki, Finland

## Abstract

Monosomy 7 and del(7q) (-7/-7q) are frequent chromosomal abnormalities detected in up to 10% of acute myeloid leukemia (AML) patients. Despite unfavorable treatment outcomes, no approved targeted therapies exist for patients with -7/-7q. Therefore, we aimed to identify novel therapeutic vulnerabilities in AML with -7/-7q. Through an analysis of data from ex vivo drug screens in 270 primary AML samples, we discovered that -7/-7q AML cells are highly sensitive to the inhibition of nicotinamide phosphoribosyltransferase (NAMPT). NAMPT is a rate-limiting enzyme in the NAD+ salvage pathway. Mechanistically, the *NAMPT* gene is located at 7q22.1, and deletion of one copy due to -7/-7q results in *NAMPT* haploinsufficiency. This leads to reduced gene expression and a therapeutically targetable vulnerability to the inhibition of NAMPT. Our results show that in -7/-7q AML, differentiated CD34+CD38+ progenitor cells are the most sensitive to the inhibition of NAMPT. In addition, we found that the combination of BCL2 inhibitor venetoclax and a NAMPT inhibitor efficiently eradicated undifferentiated AML blasts with -7/-7q. In conclusion, our findings demonstrate that AML samples with -7/-7q are highly sensitive to NAMPT inhibition, suggesting that NAMPT inhibitors have the potential to be an effective targeted therapy for patients with monosomy 7 or del(7q).

**KEY POINTS:**

- Monosomy 7 and del(7q) result in a one-copy deletion of the NAMPT gene at 7q22.1
- NAMPT haploinsufficiency causes a vulnerability to the inhibition of NAMPT

## INTRODUCTION

Monosomy 7 and deletion of chromosome arm 7q (-7/-7q) are frequent chromosomal abnormalities in up to 10% of patients with acute myeloid leukemia (AML).^1^ Monosomy 7 and del(7q) are recurrent in myeloid malignancies in both adults and children, including in juvenile myelomonocytic leukemia, ^2, 3^ myelodysplastic syndromes (MDS),^4^ and chronic myeloid leukemia. ^5^ In AML, monosomy 7 is correlated with a poor prognosis and resistance to chemotherapy, while the prognostic significance of del(7q) is less clear. This difference can be due to differences in the size of the deletion or co-occurring abnormalities.^6^ Monosomy 7 and del(7q) are among the most common chromosomal abnormalities in high-risk AML patients. Monosomy 7 often co-occurs with other markers of adverse prognosis, such as *TP53* mutations and inv(3), worsening clinical outcomes.^1^ A higher frequency of -7/-7q is observed in therapy-related AML and MDS cases. As many as 50% of therapy-related cases have -7/-7q, suggesting that chemotherapy and radiation select cells with these abnormalities.^7, 8^

Monosomy 7 and -7q result in the loss of one copy of chromosome arm 7q. The homozygous loss of both copies of 7q has not been observed, implying that the deletion of two copies of genes in 7q reduces tumor cell viability.^6^ Monosomy 7 and del(7q) lead to the haploinsufficiency of hundreds of genes. Tumor suppressor genes located on the chromosome arm 7q, which have been shown to contribute to the development of myeloid malignancies include *SAMD9*, *SAMD9L*,^9^ *KMT5C* (*MLL3*),^10, 11^ *EZH2,*^12, 13^ and *CUX1*.^14, 15^ Despite the unfavorable treatment outcome for AML patients with -7/-7q, no targeted therapies for this subgroup of patients have been approved.

Ex vivo drug sensitivity testing has emerged as a powerful tool for identifying therapeutically targetable vulnerabilities in several hematological malignancies.^16–18^ In this study, we aimed to identify novel therapeutic vulnerabilities in AML with -7/-7q. We analyzed the ex vivo drug sensitivities of 270 AML samples using a drug library with up to 527 approved and investigational oncology compounds. We performed validation of drug sensitivities using orthogonal assays, including multiparametric flow cytometry, which enables the determination of the sensitivity of distinct cell populations in leukemic bone marrow samples.

We hypothesized that -7/-7q would render AML cells dependent on distinct survival mechanisms that could be exploited to provide a novel and efficacious therapeutic strategy for patients with -7/-7q. Through an analysis of data from ex vivo drug screens in primary AML samples, we discovered that -7/-7q AML cells are highly sensitive to the inhibition of nicotinamide phosphoribosyltransferase (NAMPT). Mechanistically, the *NAMPT* gene is located at 7q22.1, and deletion of one copy due to -7/-7q results in haploinsufficiency. This leads to a therapeutically targetable vulnerability to the inhibition of NAMPT by reducing *NAMPT* gene expression in leukemia cells.

## MATERIALS AND METHODS

### Patient samples

Bone marrow (BM) or peripheral blood (PB) samples were collected after informed consent from patients with AML using protocols approved by the Institutional Review Board at the Helsinki University Hospital (permit numbers 239/13/03/00/2010, 303/13/03/01/2011, Helsinki University Hospital Ethics Committee) in compliance with the Declaration of Helsinki. Mononuclear cells were isolated using a Ficoll density gradient and suspended in conditioned medium.

### Ex vivo drug sensitivity testing with patient samples

We evaluated the drug sensitivity of freshly isolated mononuclear cells from AML patients to 527 small molecule inhibitors, including NAMPT inhibitor daporinad, using CellTiter-Glo and CellTox Green assays. Multiple NAMPT inhibitors were tested at increasing concentrations, and cells were left to recover in conditioned medium before being plated onto pre-drugged plates. After 72-hour incubation with the drugs, viability readouts were normalized to negative and positive controls, and a dose-response curve was fitted to the drug sensitivity data. The concentration range tested for daporinad, GMX1778, and LSN3154567 was 0.1-1000 nM, and for KPT-9274, 1-10000 nM. For each dose-response curve, a drug sensitivity score (DSS) was calculated as previously described.^19^

### Ex vivo drug sensitivity testing by multiparametric flow cytometry

Four NAMPT inhibitors (daporinad, GMX1778, KPT9274, and LSN3154567) were pre-plated on 96-well plates, and thawed cryopreserved mononuclear cells (MNCs) were added to the plates. After a 72-hour incubation period, the cells were stained and analyzed using flow cytometry to determine cell viability. The experiment was also performed to determine the sensitivity of -7/-7q AML to combinations of KPT-9274 and venetoclax in samples from three AML patients with an immature blast population. The viability of the CD34+CD38- population was determined after incubating the cells with either a single dose of KPT-9274, a single dose of venetoclax, or both.

### Identification of monosomy 7 and del(7q)

We retrieved clinical karyotype data for AML patient samples from the Finnish Hematology Registry and Biobank and the Hospital District of Helsinki, and Uusimaa Hematological Datalake. For AML samples without karyotype data, chromosomal copy number aberrations were identified through exome sequencing of bone marrow or peripheral blood and a matched skin biopsy sample.

### RNA-sequencing and gene expression analysis

We retrieved gene expression values for The Cancer Genome Atlas (TCGA) AML samples from cBioportal^20, 21^ and from AML samples RNA sequenced in-house. Single-cell gene expression profiles were determined using the 10X Genomics Chromium Single Cell 3’RNAseq platform. The data was processed using the Cell Ranger mfastq and count pipelines, aligned to the reference genome GRCh38, and analyzed using Seurat v4.3.0^22^ and R 4.2.1.

The complete description of the methods used in this study can be found in the supplemental materials accompanying this article.

## RESULTS

In total, 209 patients with AML and 13 healthy donors were included in the study. Patients with acute promyelocytic leukemia were excluded. We analyzed 270 bone marrow biopsy samples from patients with AML. Chromosomal abnormalities were identified for all AML samples, of which 21 (7.8%) had -7 or -7q. Biopsy samples taken at multiple time points were obtained from 45 patients. The clinical characteristics of the study cohort (FIMM AML) are shown in supplemental Tables 1 and 2. On average, the samples with -7/-7q AML had higher blast counts at the time of biopsy; otherwise, the clinical characteristics of patients with and without -7/-7q were comparable.

### AML samples with -7 or del(7q) are sensitive to the inhibition of NAMPT

To identify therapeutic vulnerabilities in AML with -7/-7q, we compared ex vivo drug sensitivities in AML samples with -7/-7q with samples with diploid chromosome 7. Fresh bone marrow mononuclear cells from patients with AML were incubated for 72 hours with increasing concentrations of up to 525 oncology drugs, followed by the determination of cell viability using the CellTiter-Glo luminescence assay and curve fitting to determine dose response. We found that AML samples with -7/-7q were more sensitive to the NAMPT inhibitor daporinad. A comparison of the daporinad dose-response curves of the representative samples with a median of IC50 in the two groups showed that half-maximal inhibition was achieved with a 4-nM-lower daporinad concentration in AML sample with -7/-7q compared with the AML sample with diploid chromosome 7 (Figure 1A).

**Figure 1.**
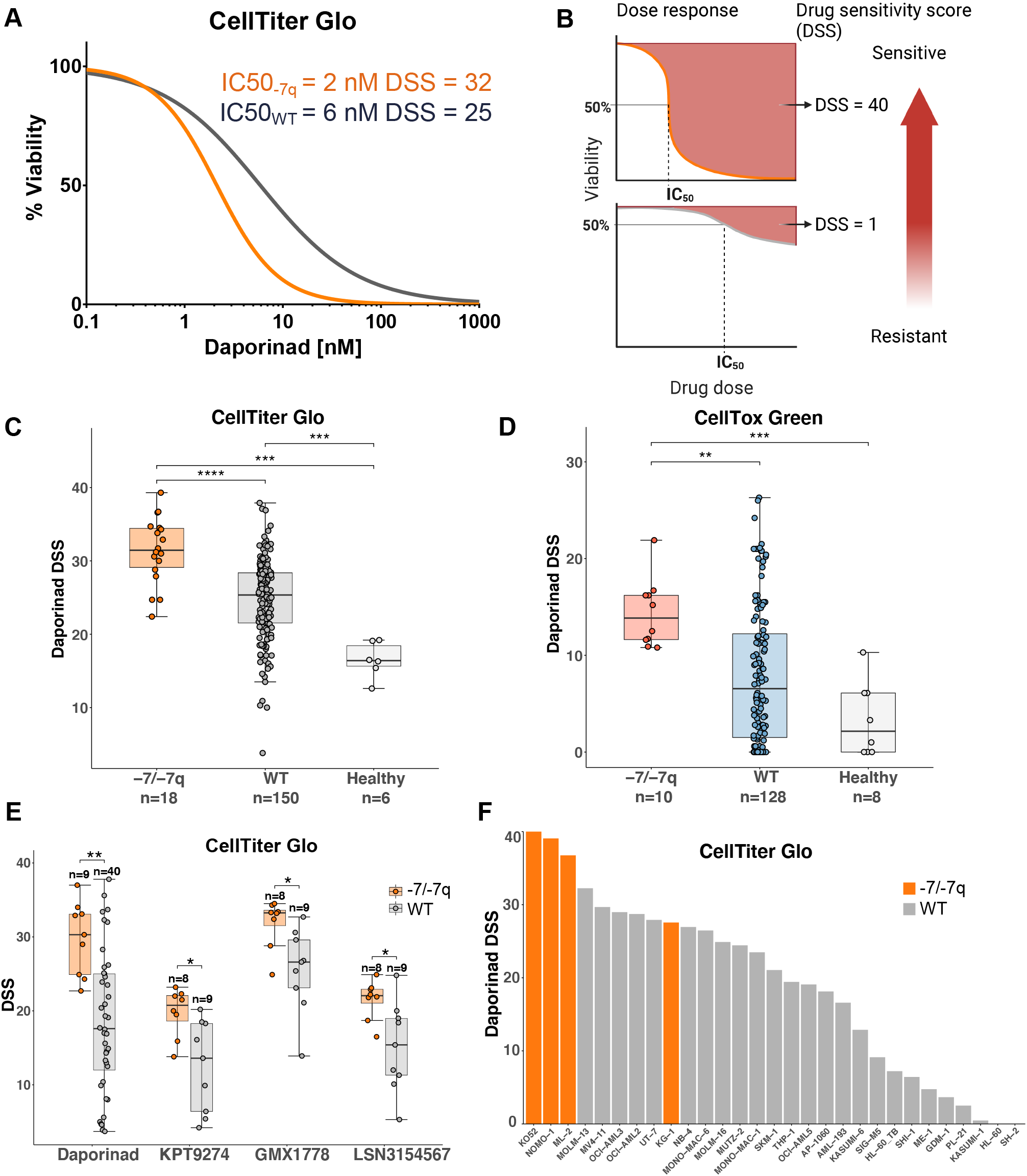
Acute myeloid leukemia (AML) cells with -7/-7q are sensitive to the inhibition of NAMPT. (A) Comparison of dose-response to NAMPT inhibitor daporinad in cells from an AML harboring del(7q) with an AML with diploid chromosome 7 (WT). Representative AML samples from -7/-7q and WT cohorts with median daporinad sensitivity are shown. Bone marrow mononuclear cells obtained from fresh biopsies were incubated with the drug for 72 hours in five concentrations, and viability was determined using the CellTiter-Glo (CTG) assay. (B) Schematic diagram illustrating the relationship of the dose-response curve and drug sensitivity score (DSS). The area delineated by a dose-response curve is transformed into a DSS using an integral function previously described.^19^ DSS values increase with increasing sensitivity to the drug being tested. (C) Comparison of daporinad sensitivity of bone marrow mononuclear cells from a cohort of patients with AML and healthy individuals. AML with -7/-7q is compared with WT AML and healthy individuals. Cell viability was measured using CTG, and (D) sensitivity to daporinad-induced cell death was measured using CellTox Green (CTxG). (E) Ex-vivo sensitivity to NAMPT inhibitors in viably-frozen cells from patients with -7/-7q AML and without (WT). (F) Sensitivity to daporinad in AML cell lines. Cell lines with -7q are shown in orange. Cells were incubated with daporinad for 72 hours, and viability was measured with CTG. *, P < 0.05; **, P < 0.01; ***, P < 0.001;****, P < 0.0001 by Mann–Whitney U test.

To facilitate the statistical analysis of drug sensitivities between the sample groups, we computed drug sensitivity scores from the dose-response curves (Figure 1B). A comparison of daporinad sensitivities in a cohort consisting of -7/-7q AML samples, AML samples with diploid chromosome 7, and healthy controls showed the viability of - 7/-7q AML samples was significantly lower after incubation with daporinad than in wild-type AML samples and samples from healthy donors (Figure 1C). Bone marrow mononuclear cells from healthy individuals were significantly more resistant to daporinad than cells from patients with AML.

To determine whether AML cells with -7/-7q were more efficiently killed by daporinad, we compared the daporinad-induced cell death in -7/-7q AML samples with wild-type AML samples using the CellTox Green cytotoxicity assay (Figure 1D). We found that daporinad killed significantly more cells in the -7/-7q AML samples than in the AML samples with diploid chromosome 7 or the samples from healthy controls.

To determine whether the reduced viability induced by NAMPT inhibitors in the -7/-7q AML samples was caused by the on-target inhibition of NAMPT, we tested the sensitivity of a cohort of AML samples to four NAMPT inhibitors with distinct chemical structures. We used viably-frozen bone marrow mononuclear cells from up to 49 patients with AML and determined the viability of the cells after incubation with each of the inhibitors using the CellTiter-Glo assay. The rationale for this experiment was that since structurally distinct NAMPT inhibitors have unique off-target activity profiles, they may exhibit activity against distinct groups of samples if the off-target effects are the dominant cause of cell killing. We found -7/-7q AML samples were significantly more sensitive than wild-type AML samples to all four NAMPT inhibitors tested, indicating that the preferential inhibition of -7/-7q AML was due to the on-target inhibition of NAMPT (Figure 1E).

Next, we examined the sensitivity of immortalized AML cell lines to NAMPT inhibition. Unlike patient cells, cell lines exhibit robust growth in vitro and represent a cell population encompassing all cell cycle stages. The sensitivity of 29 AML cell lines was tested by exposing cells to increasing concentrations of daporinad for 72 hours, followed by the determination of viability using the CellTiter-Glo assay. We found that the AML cell lines most sensitive to daporinad carried -7 or -7q (Figure 1F).

We then aimed to test can we replicate our finding using an orthogonal assay. For this purpose, we performed a dose-response evaluation using four NAMPT inhibitors (daporinad, KPT9274, GMX1778, LSN3154567) with multiparametric flow cytometry readout in viably-frozen bone marrow mononuclear cells from a cohort of AML patients. In the drug-sensitivity testing employing multiparametric flow cytometry, living cells are quantified after incubation with the drugs, whereas the CellTiter-Glo is a metabolic assay that measures the ATP released by viable cells. We tested the sensitivity of 23 cryopreserved primary AML cells with (n = 8) and without (n = 15) -7/-7q to four NAMPT inhibitors using a flow cytometry–based drug sensitivity assay. Dimly positive CD45 expression and low side scatter were used to identify leukemic blasts. Analysis of the representative samples with median daporinad IC50 showed that the AML blast cell in the sample with -7/-7q had an IC50 value of 1.5 nM, which was 16 times lower than the 24.6 nM IC50 value observed in the wild-type AML blast cells (Figure 2A). We found that the blast cells in -7/-7q AML samples were significantly more sensitive to all four NAMPT inhibitors tested than blasts from wild-type AML (Figures 2B–E). These results provided an orthogonal validation of our prior findings. Together, our results show that - 7/-7q is associated with sensitivity to NAMPT inhibition in AML.

**Figure 2.**
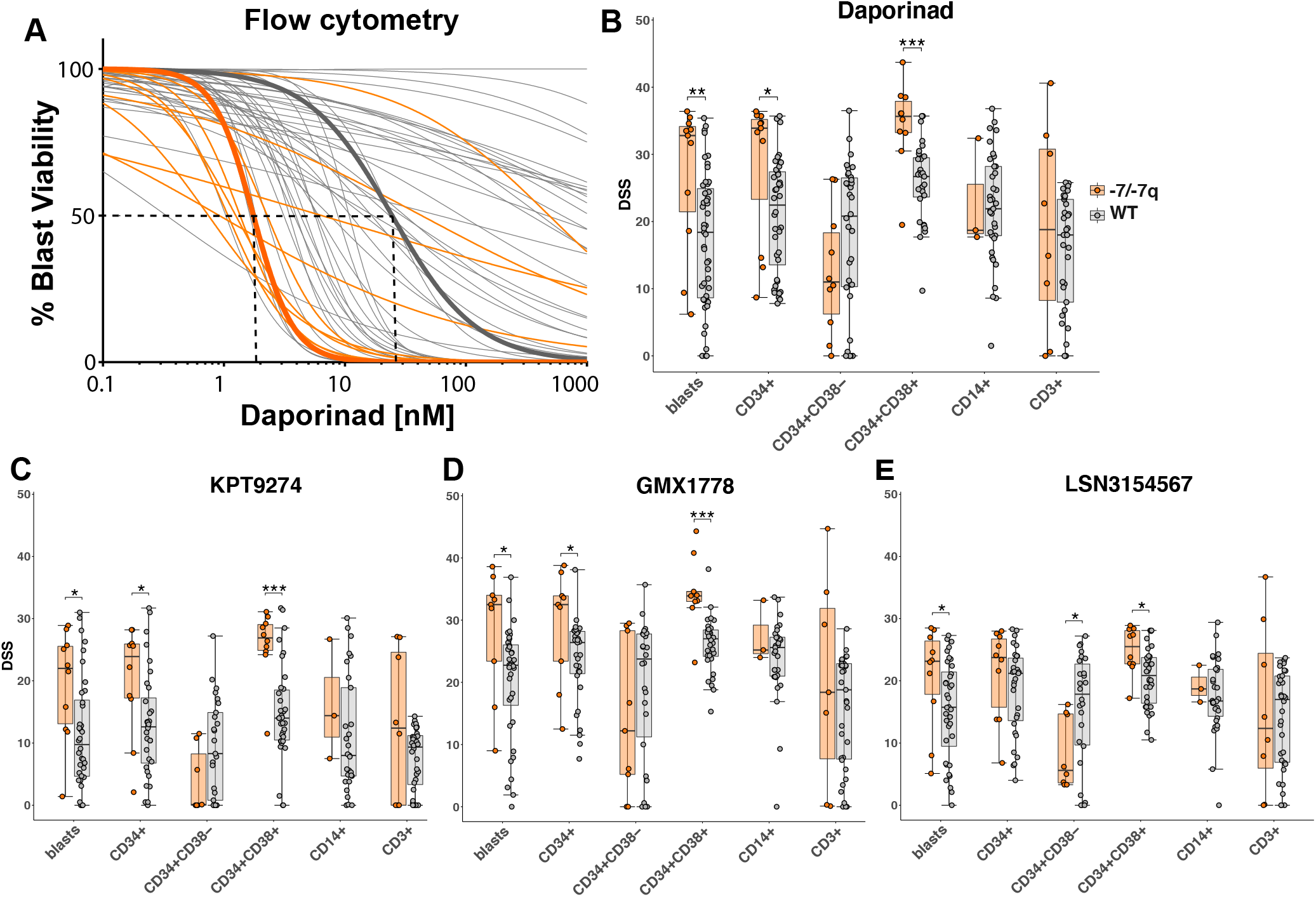
NAMPT inhibitor sensitivity of bone marrow cell populations in AML with -7/-7q. (A) Daporinad dose response of leukemic blast cells from the bone marrow of patients with AML, measured by multiparametric flow cytometry. Samples were incubated with inhibitors for 72 hours, followed by high-throughput flow cytometry–based cell viability readout. The curves highlighted in bold show patient samples with median IC50 for AML with -7/-7q (orange) and AML with diploid chromosome 7 (grey). (B) Daporinad sensitivity of cell populations in AML with -7/-7q (n = 11) compared with AML without the deletion (WT) (n = 47). (C) KPT9274 sensitivity in AML with -7/-7q (n = 10) vs. WT (n = 36). (D) GMX1778 sensitivity -7/-7q (n = 9) vs. WT (n = 36). (E) LSN3154567 sensitivity in -7/-7q (n = 10) vs. WT (n = 36).

### Sensitivity of bone marrow cell populations to NAMPT inhibition in -7/-7q AML

To determine the effect of the inhibition of NAMPT on blast, progenitor, and lymphocyte cell populations from the bone marrow of patients with -7/-7q AML, we analyzed the sensitivity of cryopreserved patient cells against daporinad, KPT9274, GMX1778, and LSN3154567 using multiparametric flow cytometry. The flow antibody panel (supplemental Table 3) consisted of viability and cell surface markers that allowed us to distinguish the major viable cell populations in the AML MNCs. The markers were used as follows: CD34 for immature stem and progenitor cells, CD38 for more-differentiated myeloid cells, CD14 for (pro-) monocytes, and CD3 for lymphocytes. Dimly positive CD45 expression and low side scatter were used to identify leukemic blasts. We found that more-differentiated myeloblasts identified by CD34 and CD38 expression were significantly more sensitive to NAMPT inhibition in -7/-7q than in wild-type AML. In contrast, less-differentiated CD34+CD38- myeloblasts from -7/-7q AML did not exhibit increased sensitivity to NAMPT inhibition. We did not observe a significant difference between the sensitivity of differentiated monocytes or lymphocytes in -7/-7q AML and AML lacking the deletion. Lymphocytes from -7/-7q AML showed notably large variability in responses to NAMPT inhibition (Figures 2B–E). To determine the relative potency of NAMPT inhibitors, we analyzed the drug sensitivity score ranges. Compared to the other inhibitors, GMX1778 seemed to be the most potent drug against different cell types, consistently returning higher DSS scores. Overall, our results show that in - 7/-7q AML, differentiated progenitor cells are the most sensitive to the inhibition of NAMPT.

### AML with del(7q) that does not encompass the *NAMPT* gene locus lack sensitivity to NAMPT inhibition

The *NAMPT* locus remains diploid in cases where the del(7q) breakpoint occurs on the telomeric side of the gene. We sought to determine whether the location of the del(7q) breakpoint influences drug sensitivity. To identify breakpoint locations in the AML samples with del(7q), we analyzed copy number profiles defined by exome sequencing in 19 del(7q) AML. We found that, in two patients, the breakpoint was on the telomeric side of the NAMPT locus, maintaining diploid *NAMPT* (Figure 3A). We evaluated the effect of daporinad on viability by CellTiter-Glo (Figure 3B), and cell killing using CellTox Green assays (Figure 3C) in del(7q) AML with diploid (*NAMPT^+/+^*) and -7/-7q AML with haploid *NAMPT* (*NAMPT^+/-^*). This revealed that *NAMPT*^+/+^ AML samples lacked sensitivity to daporinad, while *NAMPT*^+/-^ samples were highly sensitive (Figure 3B, C, and supplemental Figure 1). Our results show del(7q) AML are sensitive to the inhibition of NAMPT when the deletion breakpoint is centromeric to the *NAMPT* locus at 7q22.3, leading to *NAMPT* haploinsufficiency. In comparison, AML with the del(7q) breakpoint on the telomeric side of the *NAMPT* locus do not exhibit increased sensitivity to NAMPT inhibitors.

**Figure 3.**
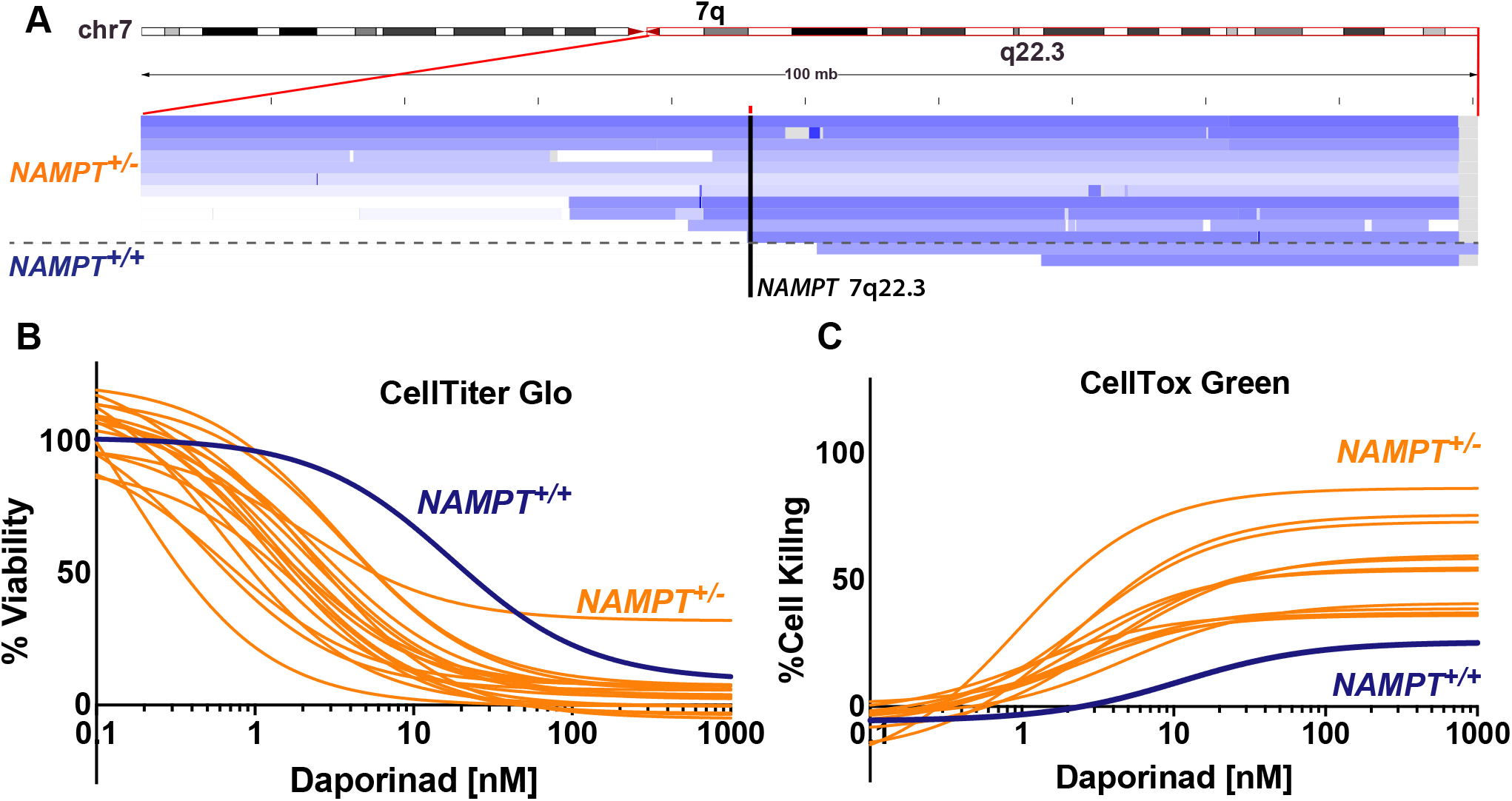
AML with 7q deletion that does not encompass *NAMPT* lacks sensitivity to NAMPT inhibition. (A) Genomic coverage plot showing the location of the deletion breakpoint in -7q AML samples with respect to the *NAMPT* gene locus. In two AML cases, a del(7q) breakpoint was on the telomeric side of *NAMPT*, maintaining the locus diploid (*NAMPT^+/+^*). The deletion breakpoint locations were determined based on exome sequencing. (B) Ex-vivo daporinad dose response of a *NAMPT*^+/+^ AML sample with -7q compared with -AML samples with *NAMPT* haploinsufficiency (*NAMPT*^+/-^). Cell viability was measured by CTG and (B), and cell death using CTxG (C).

### AML samples with -7/-7q have low *NAMPT* expression levels

Monosomy 7 and del(7q) lead to the deletion of one copy of the *NAMPT* gene in most cases (Figure 4A). Haploinsufficient gene expression is characterized by a reduced ability of cells to upregulate RNA expression. To determine whether *NAMPT* exhibits haploinsufficient gene expression in AML with -7/-7q, we analyzed *NAMPT* gene expression data from two AML cohorts. We found that the dynamic range of *NAMPT* expression was reduced in samples with -7/-7q, both in TCGA AML cohort (Figure 4B) and our in-house cohort of AML samples from Finland (Figure 4C). These results indicate that *NAMPT* exhibits haploinsufficient gene expression in AML with -7/-7q.

**Figure 4.**
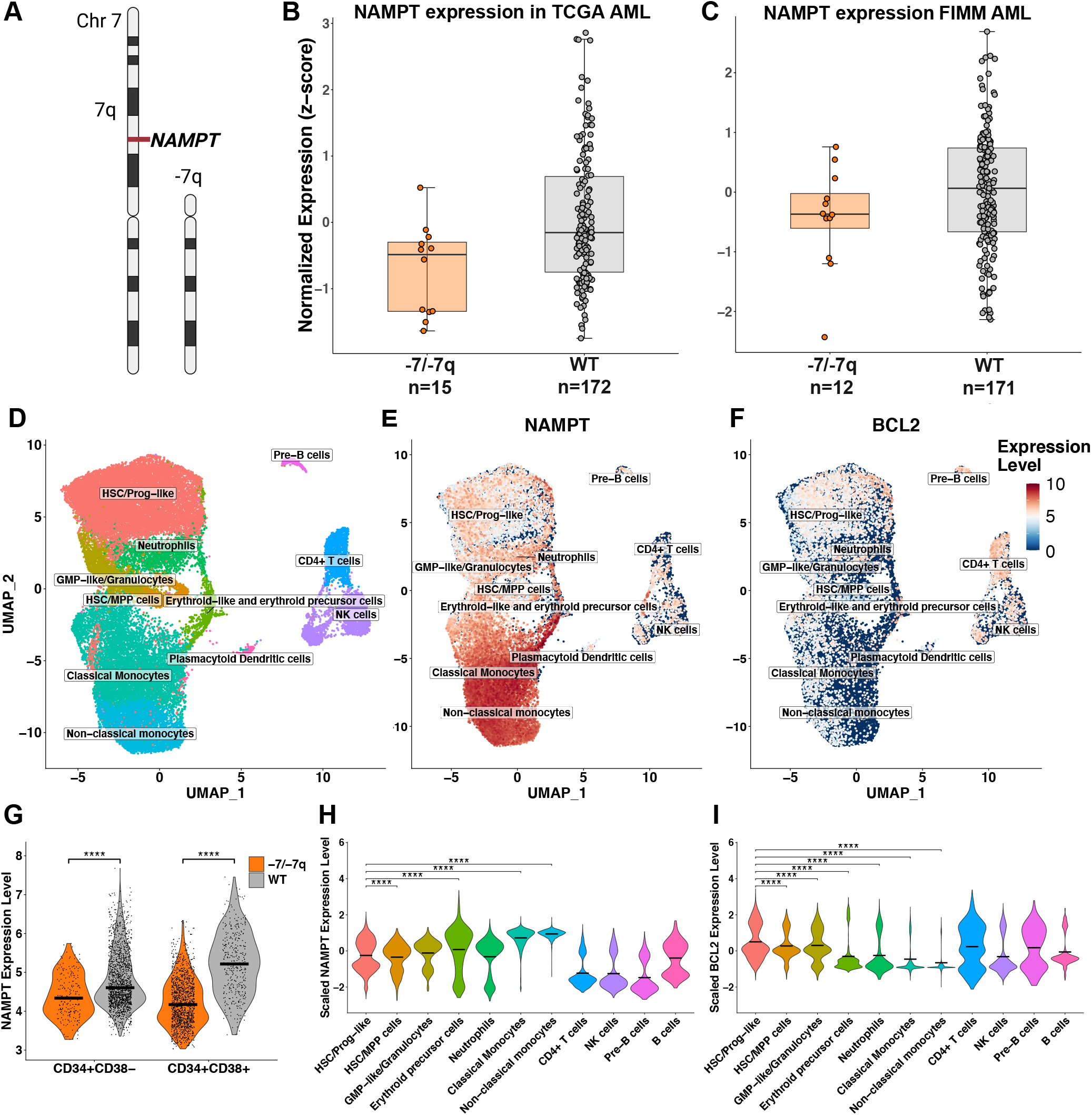
AML with -7/-7q have reduced *NAMPT* expression. (A) Diagram illustrating the deletion of one copy of *NAMPT* locus resulting from del(7q). Comparison of the level of *NAMPT* gene expression in the bone marrow mononuclear cells from AML with -7/-7q compared with AML lacking the deletion (WT). Gene expression was determined by whole transcriptome RNA sequencing. *NAMPT* gene expression analysis was performed on independent AML patient cohorts from TCGA (B) and the Institute for Molecular Medicine Finland (FIMM) (C). (D) Uniform Manifold Approximation and Projection (UMAP) representation of the different cell types identified from single-cell RNA sequencing data from eight AML patient samples. The group consists of one sample with monosomy 7 and seven samples that were wild-type. Cells are colored based on clusters identified using ScType and in-house gene sets. (E) UMAP representation of log-normalized *NAMPT* gene expression level across the total cell cohort. *NAMPT* expression increases towards the differentiated myeloid cell types. (F) UMAP representation of log-normalized *BCL2* gene expression. While *BCL2* is expressed in all different cell types, cells that belong to the HSC/progenitor-like cell cluster show the most expression in the myeloid cells. (G) Violin plot showing log-normalized *NAMPT* gene expression in CD34+CD38- and CD34+CD38+ cells. Cells were selected from the total cell cohort based on their CD34 and CD38 expression levels. *NAMPT* expression is significantly lower in cells that carry monosomy 7 compared to cells that are considered WT. (H) Violin plot showing scaled *NAMPT* gene expression in the 11 different cell clusters. *NAMPT* expression is highest in the more-differentiated myeloid cells (erythroid-like, neutrophil-like, and monocyte-like cells) compared to myeloid progenitor or lymphoid cells. (I) Violin plot showing scaled *BCL2* gene expression in cell clusters. *BCL2* is expressed at a significantly higher level in the HSC/progenitor-like cells compared to other myeloid cell clusters. *, P < 0.05; **, P < 0.01; ***, P < 0.001; ****, P < 0.0001 by Dunn’s test following a Kruskal-Wallis H test.

### Differentiated CD34+CD38+ AML myeloblasts have high *NAMPT* expression

Since we found that more-differentiated bone marrow myeloid progenitor cells from AML patients were more sensitive to the inhibition of NAMPT than their less-differentiated counterparts, we sought to determine whether these cells expressed higher levels of *NAMPT*. To determine *NAMPT* expression levels in myeloid cell populations in the bone marrow of patients with AML, we analyzed viably-frozen mononuclear cells using single-cell RNA sequencing from eight patients, of which one had -7 (Figure 4D). We found that more-differentiated myeloid cells had significantly higher levels of *NAMPT* expression than primitive myeloid cells and hematopoietic stem cells (Figures 4E and 4H). To examine similar cell populations as we did using multiparametric flow cytometry–based drug sensitivity testing, we selected a subpopulation of cells based on their *CD34* and *CD38* gene expression to represent CD34+CD38- and CD34+CD38+ cells. In these cells, we found that *NAMPT* gene expression was significantly lower in AML cells with -7 than in WT cells (Figure 4G), supporting our findings from the bulk RNA sequencing studies. In addition, CD34+CD38+ cells in the WT samples also had significantly higher expression of *NAMPT* than CD34+CD38- cells, indicating that CD34+CD38+ cells are likely to be more reliant on NAMPT for their metabolism and, therefore, more sensitive to NAMPT inhibition.

Since HSC-like blasts (CD34+CD38-) are less sensitive to NAMPT inhibitors, other vulnerabilities must be exploited to inhibit these cells. HSC-like blasts are known to be more reliant on BCL2 for their survival.^23^ We studied the level of *BCL2* expression in the myeloid cell clusters since BCL2 could be targeted with BCL2 inhibitors in combination with NAMPT inhibitors (Figure 4F). In the myeloid cells, *BCL2* was expressed most in the HSC/progenitor-like cells, and the expression decreased significantly toward the more-differentiated cell states (Figure 4I), while the opposite was true for *NAMPT* (Figure 4H). These results suggest a combination with inhibitors of BCL2 could help to eradicate the stem cell–like blasts that are less dependent on NAMPT but depend on BCL2 for survival.

### Combination of BCL2 inhibitor venetoclax and a NAMPT inhibitor efficiently eradicated undifferentiated AML blasts with -7/-7q

We hypothesized that less-differentiated cells could be inhibited by combining a NAMPT inhibitor with a BCL2-specific inhibitor. To test this, we incubated samples from three patients with AML harboring -7/-7q and a quantifiable CD34+CD38- cell population with either the NAMPT inhibitor KPT-9274 at 100 nM alone, the BCL2 specific inhibitor venetoclax at 30 nM alone, or a combination of both agents for 72 hours. Afterward, we quantified the cell death caused by the treatments within the undifferentiated CD34+CD38- cell population in each of the samples by multiparameter flow cytometry (Figure 5). As expected, while KPT-9274 caused limited cell killing of CD34+CD38- blasts in the samples, venetoclax resulted in higher toxicity in these cells. Notably, the combination resulted in near total cell death of the immature blasts in the samples, indicating that the dual inhibition of NAMPT and BCL2 represents an effective strategy for targeting less-differentiated cells with -7/-7q.

**Figure 5.**
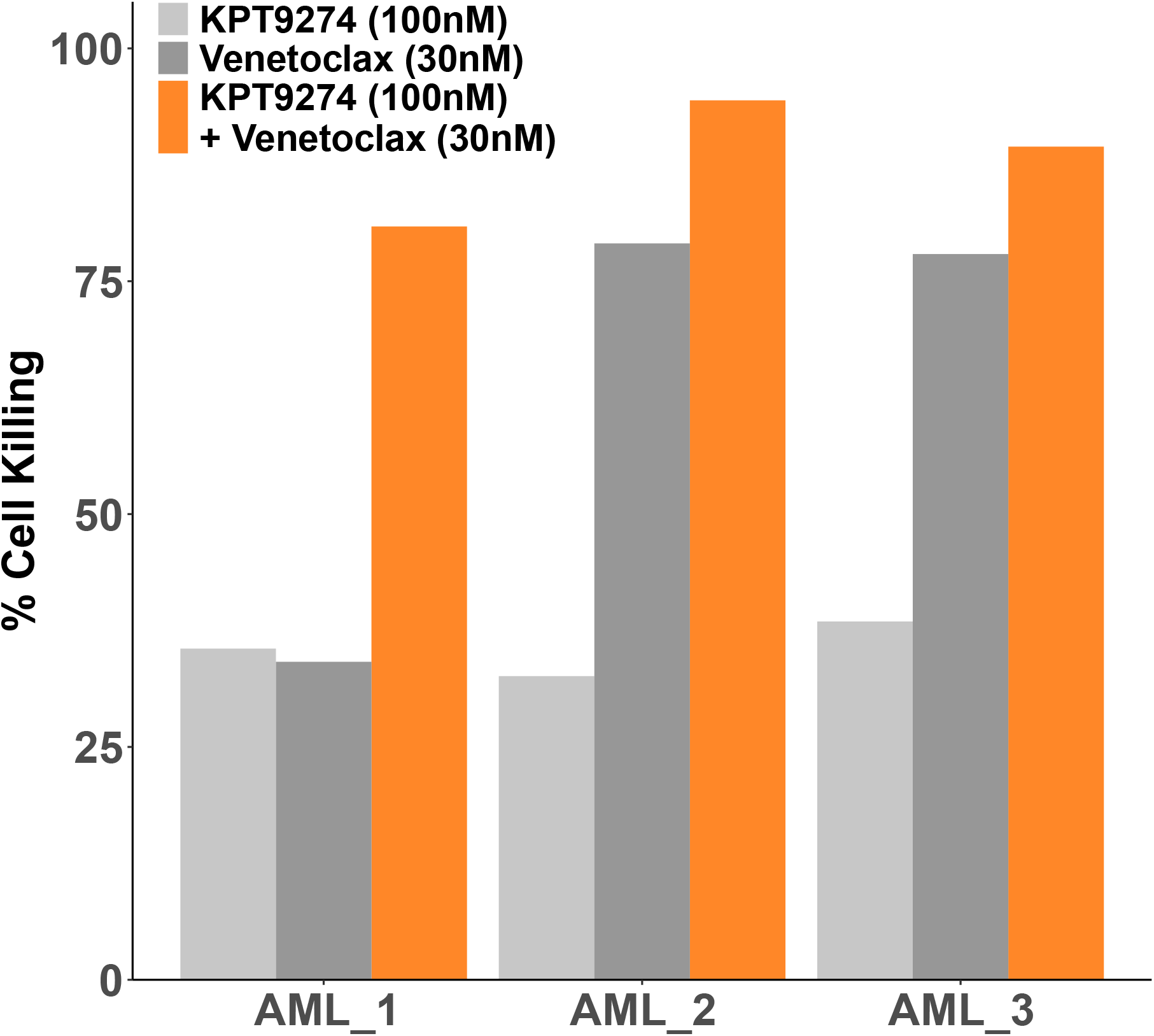
A combination of NAMPT and BCL2 inhibition effectively kills immature blasts. Barplot showing the percentage of cell death within the immature (CD34+CD38-) blast population in samples from three different AML patients with -7/-7q as measured by multiparametric flow cytometry following a 72 h incubation with NAMPT inhibitor KPT- 9274 at 100 nM alone (light grey), BCL2 inhibitor venetoclax at 30 nM alone (dark grey), or both agents (orange).

## DISCUSSION

In this study, we identified NAMPT inhibition as a novel therapeutically targetable vulnerability in AML cells with -7/-7q. We analyzed the ex vivo drug sensitivities of primary AML patient samples in the FIMM AML cohort^18, 24^ that were based on viability and cytotoxicity assays. As a result, we found that samples with -7/-7q were more sensitive to NAMPT inhibition compared to wild-type AML and healthy samples. Additional experiments using structurally distinct NAMPT inhibitors confirmed that the sensitivity of -7/-7q AML samples to NAMPT inhibitors was due to on-target inhibition.

Cancer cells are known to have high metabolic activity and increased nicotinamide adenine dinucleotide (NAD+) turnover compared to normal cells because of the activity of enzymes such as sirtuins and poly (ADP-ribose) and polymerase I (PARP-1) that use NAD+ as a substrate.^25, 26^ Cancer cells do not efficiently utilize the de novo or Preiss-Handler pathways for NAD+ production due to low levels of nicotinic acid and tryptophan and the lack of expression of key enzymes in these pathways. Consequently, cancer cells generate NAD+ mainly through the NAD+ salvage pathway.^27^ NAMPT is the rate-limiting enzyme in the NAD+-salvage pathway and has been extensively researched as a potential drug target for targeting cancer cell metabolism.^28^ By inhibiting NAMPT, the salvage pathway is halted, resulting in a drop in NAD+ levels, followed by ATP depletion and, eventually, death of cancer cells.^27^ Our results also show a higher dependence of AML cells on NAMPT compared to healthy cells, as demonstrated by their significantly higher sensitivity to NAMPT inhibition (Figures 1C, D).

Several NAMPT inhibitors have been developed and have shown encouraging results in terms of anticancer efficacy in vitro and in vivo models of different solid^29–32^ and hematological malignancies.^33^ These inhibitors include daporinad (APO866, FK866),^34^ GMX1778 (CHS 828),^35^ LSN3154567,^36^ KPT9274,^37^ and most recently, OT82.^38^ However, when evaluated in phase I/II clinical trials, NAMPT inhibitors have been reported to have limited drug efficacy profiles and severe toxic side effects, including thrombocytopenia, anemia, and gastrointestinal symptoms.^39–43^ Studies on rodent models have also shown that NAMPT inhibition causes retinopathy.^44^ Since dose-limiting toxicities have so far prevented NAMPT inhibitors from achieving desirable therapeutic effects and clinical approval, our identification of AML with -7/-7q as a particularly responsive patient population could improve the clinical profile of NAMPT inhibitors, notably by utilizing an actionable therapeutic window that limits side effects while maintaining a high level of efficacy.

Hypothesizing that the observed sensitivity of AML with -7/-7q to NAMPT inhibitors is due to the loss of one copy of the *NAMPT* gene, termed *NAMPT* haploinsufficiency, we investigated the role of the location of the del(7q) breakpoint on drug sensitivity in AML. By analyzing copy number profiles defined by exome sequencing in del(7q) AML samples, we found that in two patients, the del(7q) breakpoint was on the telomeric side of the *NAMPT* locus, maintaining diploid *NAMPT*. These samples did not show the increased sensitivity to daporinad observed in samples with *NAMPT* haploinsufficiency, and their response was comparable to wild-type AML samples. These results show del(7q) AML are sensitive to the inhibition of NAMPT when the deletion breakpoint is located on the centromeric side of the *NAMPT* locus at 7q22.3, causing *NAMPT* haploinsufficiency. Therefore, a precise characterization of del(7q) is necessary to identify the patients who can benefit from NAMPT-targeted therapy. Conventional cytogenetic analysis and fluorescence in situ hybridization are used routinely in the diagnosis of acute myeloid leukemia.^45^ However, these methods lack the resolution to detect the 7q breakpoint location accurately. The breakpoint location can be determined using high-resolution methods, such as whole-genome sequencing.^46^

More differentiated myeloid cells have high *NAMPT* expression and are sensitive to NAMPT inhibition. We investigated the effect of NAMPT inhibition on different cell populations in the bone marrow of patients with -7/-7q AML using a flow cytometry-based drug sensitivity assay. We found that the more differentiated myeloblasts were significantly more sensitive to NAMPT inhibition in -7/-7q than in wild-type AML. However, less differentiated myeloblasts from -7/-7q AML did not exhibit increased sensitivity to NAMPT inhibition. Overall, our results suggest that differentiated progenitor cells are the most sensitive cell population to NAMPT inhibition in -7/-7q AML. Furthermore, our single-cell gene expression analysis of primary AML samples confirmed that highly differentiated myeloblasts express high levels of *NAMPT*. Taken together, these results suggest mature myeloid cells rely heavily on NAMPT for survival, making them more vulnerable to NAMPT inhibition.

The results of our study indicate that targeting both NAMPT and BCL2 may represent a promising approach for treating AML patients with -7/-7q chromosomal abnormalities. Our flow cytometry-based drug sensitivity assay revealed that undifferentiated (CD34+CD38-) malignant cells in AML samples with -7/-7q are less responsive to NAMPT inhibition alone compared to more mature (CD34+CD38+) blasts. However, our experiments combining a NAMPT inhibitor (KPT-9274) with a BCL2-specific inhibitor (venetoclax) show that this dual therapy resulted in near-total cell death of undifferentiated blasts in these samples. These results support the idea that co-targeting the NAMPT and BCL2 pathways may be a more effective strategy for eliminating undifferentiated AML cells with -7/-7q chromosomal abnormalities. Additional studies are needed to validate these findings in larger patient populations and to further investigate the safety and efficacy of this dual-therapy approach.

Our results identify NAMPT inhibition as a novel therapeutic vulnerability in AML cells with -7/-7q chromosomal abnormalities. Instances of chromosomal abnormality conferring increased susceptibility to a particular treatment have been previously described, notably in MDS. Specifically, patients with MDS carrying -5q have shown exceptional sensitivity to the immunomodulatory drug lenalidomide. This observation was supported by several studies conducted in vitro and in vivo, as well as in clinical trials, ultimately resulting in approval of the drug for clinical use in these patients.^47^

In conclusion, our findings demonstrate that AML cells with -7/-7q are highly sensitive to NAMPT inhibition, suggesting that NAMPT inhibitors have the potential to be an effective targeted therapy for AML with -7/-7q. Additionally, our results propose NAMPT inhibitors as a suitable combination partner for venetoclax for -7/-7q patients. Further research is needed to investigate the clinical potential of NAMPT inhibitors in the treatment of AML patients with -7/-7q. In addition, as monosomy 7 and deletion 7q are observed in a number of other blood disorders, including myelodysplastic syndromes, the efficacy of NAMPT inhibitors is worth evaluating in these patients.

## AUTHORSHIP

S.E., C.A.H. conceived the project and provided leadership. S.E., J.S., and N.I. analyzed and interpreted the data. DM provided the cell line data. M.K., B.T.G., K.P. collected the samples and interpreted clinical data. S.E., J.S., N.I., D.M., and C.A.H. wrote the paper. All authors critically read the paper, provided constructive comments, and agreed to the content.

Conflict-of-interest disclosure: BTG has served as a consultant for BerGenBio and Pfizer Inc.; holds stock options in privately-held companies Alden Cancer Therapy and KinN Therapeutics; has provided consultancy services to Novartis. MK is an advisor for Novartis, Faron Pharmaceuticals, Bristol-Myers Squibb, Pfizer, and AbbVie; has received consultancy fees, research funding, and speaker’s bureau involvement from AbbVie, as well as consultancy fees from Astellas Pharma. KP has been awarded honoraria from Pfizer, Novartis, Incyte, Bristol-Myers Squibb, Astellas, and AbbVie; has received research funding from Celgene/Bristol-Myers Squibb, Incyte, Pfizer, and Novartis. CAH has received research funding from Oncopeptides, IMI2 projects HARMONY and HARMONY PLUS, WntResearch, Orion, Kronos Bio, Novartis, Celgene, Zentalis Pharmaceuticals, and Amgen; has been awarded honoraria from Amgen. The remaining author declare no competing financial interests.

Correspondence: Caroline Heckman, Institute for Molecular Medicine Finland, University of Helsinki, FI-00290, and Samuli Eldfors, Massachusetts General Hospital Cancer Center, Charlestown, MA 02129.

## Data Availability

Drug sensitivity and processed gene expression data from the FIMM AML cohort used in the study are available at

https://doi.org/10.5281/zenodo.7274740

## ACKNOWLEDGEMENTS

We thank the Helsinki Biobank for the control samples and the Finnish Hematology Registry and Clinical Biobank (FHRB) for providing samples. Drug plate preparation was carried out at the FIMM High Throughput Biomedicine Unit, and single-cell sequencing at the FIMM Single-Cell Analytics, which are hosted by the University of Helsinki and supported by HiLIFE and Biocenter Finland. J.S. and N.I. are Ph.D. candidates at the University of Helsinki. This work is submitted in partial fulfillment of the requirement for the Ph.D.

This work was supported by the University of Helsinki, Cancer Foundation Finland, the Academy of Finland (grants 334781, 1320185) (C.A.H.), (grant 334273), the Sigrid Jusélius Foundation and the Magnus Ehrnrooth Foundation (S.E.).

## REFERENCES

1. Papaemmanuil E, Gerstung M, Bullinger L, et al. Genomic Classification and Prognosis in Acute Myeloid Leukemia. New England Journal of Medicine. 2016;374(23):2209–2221.

2. Locatelli F, Crotta A, Ruggeri A, et al. Analysis of risk factors influencing outcomes after cord blood transplantation in children with juvenile myelomonocytic leukemia: a EUROCORD, EBMT, EWOG-MDS, CIBMTR study. Blood. 2013;122(12):2135– 2141.

3. Hasle H, Alonzo TA, Auvrignon A, et al. Monosomy 7 and deletion 7q in children and adolescents with acute myeloid leukemia: an international retrospective study. Blood. 2007;109(11):4641–4647.

4. Haase D, Germing U, Schanz J, et al. New insights into the prognostic impact of the karyotype in MDS and correlation with subtypes: evidence from a core dataset of 2124 patients. Blood. 2007;110(13):4385–4395.

5. Johansson B, Fioretos T, Mitelman F. Cytogenetic and Molecular Genetic Evolution of Chronic Myeloid Leukemia. Acta Haematol. 2002;107(2):76–94.

6. Inaba T, Honda H, Matsui H. The enigma of monosomy 7. Blood. 2018;131(26):2891–2898.

7. McNerney ME, Godley LA, Le Beau MM. Therapy-related myeloid neoplasms: when genetics and environment collide. Nat Rev Cancer. 2017;17(9):513–527.

8. Smith SM, Le Beau MM, Huo D, et al. Clinical-cytogenetic associations in 306 patients with therapy-related myelodysplasia and myeloid leukemia: the University of Chicago series. Blood. 2003;102(1):43–52.

9. Nagamachi A, Matsui H, Asou H, et al. Haploinsufficiency of SAMD9L, an Endosome Fusion Facilitator, Causes Myeloid Malignancies in Mice Mimicking Human Diseases with Monosomy 7. Cancer Cell. 2013;24(3):305–317.

10. Chen C, Liu Y, Rappaport AR, et al. MLL3 Is a Haploinsufficient 7q Tumor Suppressor in Acute Myeloid Leukemia. Cancer Cell. 2014;25(5):652–665.

11. Chen R, Okeyo-Owuor T, Patel RM, et al. Kmt2c mutations enhance HSC self-renewal capacity and convey a selective advantage after chemotherapy. Cell Reports. 2021;34(7):108751.

12. Muto T, Sashida G, Oshima M, et al. Concurrent loss of Ezh2 and Tet2 cooperates in the pathogenesis of myelodysplastic disorders. Journal of Experimental Medicine. 2013;210(12):2627–2639.

13. Sashida G, Harada H, Matsui H, et al. Ezh2 loss promotes development of myelodysplastic syndrome but attenuates its predisposition to leukaemic transformation. Nat Commun. 2014;5(1):4177.

14. McNerney ME, Brown CD, Wang X, et al. CUX1 is a haploinsufficient tumor suppressor gene on chromosome 7 frequently inactivated in acute myeloid leukemia. Blood. 2013;121(6):975–983.

15. Wong CC, Martincorena I, Rust AG, et al. Inactivating CUX1 mutations promote tumorigenesis. Nat Genet. 2014;46(1):33–38.

16. Eldfors S, Kuusanmäki H, Kontro M, et al. Idelalisib sensitivity and mechanisms of disease progression in relapsed TCF3-PBX1 acute lymphoblastic leukemia. Leukemia. 2017;31(1):51–57.

17. Kuusanmäki H, Dufva O, Vähä-Koskela M, et al. Erythroid/megakaryocytic differentiation confers BCL-XL dependency and venetoclax resistance in acute myeloid leukemia. Blood. 2022;blood.2021011094.

18. Malani D, Kumar A, Brück O, et al. Implementing a Functional Precision Medicine Tumor Board for Acute Myeloid Leukemia. Cancer Discovery. 2022;12(2):388–401.

19. Yadav B, Pemovska T, Szwajda A, et al. Quantitative scoring of differential drug sensitivity for individually optimized anticancer therapies. Sci Rep. 2014;4:5193.

20. Cancer Genome Atlas Research Network, Ley TJ, Miller C, et al. Genomic and epigenomic landscapes of adult de novo acute myeloid leukemia. N Engl J Med. 2013;368(22):2059–2074.

21. Gao J, Aksoy BA, Dogrusoz U, et al. Integrative Analysis of Complex Cancer Genomics and Clinical Profiles Using the cBioPortal. Science Signaling. 2013;6(269):pl1–pl1.

22. Hao Y, Hao S, Andersen-Nissen E, et al. Integrated analysis of multimodal single-cell data. Cell. 2021;184(13):3573–3587.e29.

23. Waclawiczek A, Leppä A-M, Renders S, et al. Combinatorial BCL-2 family expression in Acute Myeloid Leukemia Stem Cells predicts clinical response to Azacitidine/Venetoclax. Cancer Discovery. 2023;CD-22-0939.

24. Pemovska T, Kontro M, Yadav B, et al. Individualized Systems Medicine Strategy to Tailor Treatments for Patients with Chemorefractory Acute Myeloid Leukemia. Cancer Discov. 2013;3(12):1416–1429.

25. Epstein T, Gatenby RA, Brown JS. The Warburg effect as an adaptation of cancer cells to rapid fluctuations in energy demand. PLOS ONE. 2017;12(9):e0185085.

26. Zhu Y, Liu J, Park J, Rai P, Zhai RG. Subcellular compartmentalization of NAD+ and its role in cancer: A sereNADe of metabolic melodies. Pharmacology & Therapeutics. 2019;200:27–41.

27. Xiao Y, Elkins K, Durieux JK, et al. Dependence of Tumor Cell Lines and Patient-Derived Tumors on the NAD Salvage Pathway Renders Them Sensitive to NAMPT Inhibition with GNE-618. Neoplasia. 2013;15(10):1151-IN23.

28. Sampath D, Zabka TS, Misner DL, O’Brien T, Dragovich PS. Inhibition of nicotinamide phosphoribosyltransferase (NAMPT) as a therapeutic strategy in cancer. Pharmacology & Therapeutics. 2015;151:16–31.

29. Kozako T, Aikawa A, Ohsugi T, et al. High expression of NAMPT in adult T-cell leukemia/lymphoma and anti-tumor activity of a NAMPT inhibitor. European Journal of Pharmacology. 2019;865:172738.

30. Nacarelli T, Fukumoto T, Zundell JA, et al. NAMPT Inhibition Suppresses Cancer Stem-like Cells Associated with Therapy-Induced Senescence in Ovarian Cancer. Cancer Research. 2020;80(4):890–900.

31. Zhang C, Tong J, Huang G. Nicotinamide Phosphoribosyl Transferase (Nampt) Is a Target of MicroRNA-26b in Colorectal Cancer Cells. PLOS ONE. 2013;8(7):e69963.

32. Zhang H, Zhang N, Liu Y, et al. Epigenetic Regulation of *NAMPT* by *NAMPT-AS* Drives Metastatic Progression in Triple-Negative Breast Cancer. Cancer Research. 2019;79(13):3347–3359.

33. Nahimana A, Attinger A, Aubry D, et al. The NAD biosynthesis inhibitor APO866 has potent antitumor activity against hematologic malignancies. Blood. 2009;113(14):3276–3286.

34. Hasmann M, Schemainda I. FK866, a Highly Specific Noncompetitive Inhibitor of Nicotinamide Phosphoribosyltransferase, Represents a Novel Mechanism for Induction of Tumor Cell Apoptosis. Cancer Research. 2003;63(21):7436–7442.

35. Watson M, Roulston A, Bélec L, et al. The small molecule GMX1778 is a potent inhibitor of NAD+ biosynthesis: strategy for enhanced therapy in nicotinic acid phosphoribosyltransferase 1-deficient tumors. Mol Cell Biol. 2009;29(21):5872– 5888.

36. Zhao G, Green CF, Hui Y-H, et al. Discovery of a Highly Selective NAMPT Inhibitor That Demonstrates Robust Efficacy and Improved Retinal Toxicity with Nicotinic Acid Coadministration. Molecular Cancer Therapeutics. 2017;16(12):2677–2688.

37. Mitchell SR, Larkin K, Grieselhuber NR, et al. Selective targeting of NAMPT by KPT- 9274 in acute myeloid leukemia. Blood Adv. 2019;3(3):242–255.

38. Korotchkina L, Kazyulkin D, Komarov PG, et al. OT-82, a novel anticancer drug candidate that targets the strong dependence of hematological malignancies on NAD biosynthesis. Leukemia. 2020;34(7):1828–1839.

39. Holen K, Saltz LB, Hollywood E, Burk K, Hanauske A-R. The pharmacokinetics, toxicities, and biologic effects of FK866, a nicotinamide adenine dinucleotide biosynthesis inhibitor. Invest New Drugs. 2008;26(1):45–51.

40. Hovstadius P, Larsson R, Jonsson E, et al. A Phase I Study of CHS 828 in Patients with Solid Tumor Malignancy1. Clinical Cancer Research. 2002;8(9):2843–2850.

41. Pishvaian MJ, Marshall JL, Hwang JH, et al. A phase 1 trial of GMX1777: An inhibitor of nicotinamide phosphoribosyl transferase (NAMPRT). JCO. 2008;26(15_suppl):14568–14568.

42. Ravaud A, Cerny T, Terret C, et al. Phase I study and pharmacokinetic of CHS-828, a guanidino-containing compound, administered orally as a single dose every 3weeks in solid tumours: An ECSG/EORTC study. European Journal of Cancer. 2005;41(5):702–707.

43. von Heideman A, Berglund Å, Larsson R, Nygren P. Safety and efficacy of NAD depleting cancer drugs: results of a phase I clinical trial of CHS 828 and overview of published data. Cancer Chemother Pharmacol. 2010;65(6):1165–1172.

44. Zabka TS, Singh J, Dhawan P, et al. Retinal Toxicity, in vivo and in vitro, Associated with Inhibition of Nicotinamide Phosphoribosyltransferase. Toxicological Sciences. 2015;144(1):163–172.

45. Döhner H, Wei AH, Appelbaum FR, et al. Diagnosis and management of AML in adults: 2022 recommendations from an international expert panel on behalf of the ELN. Blood. 2022;140(12):1345–1377.

46. Macintyre G, Ylstra B, Brenton JD. Sequencing Structural Variants in Cancer for Precision Therapeutics. Trends in Genetics. 2016;32(9):530–542.

47. Giagounidis A, Mufti GJ, Fenaux P, et al. Lenalidomide as a disease-modifying agent in patients with del(5q) myelodysplastic syndromes: linking mechanism of action to clinical outcomes. Ann Hematol. 2014;93(1):1–11.

