## Supplemental Material for "Monosomy 7/del(7q) Cause Sensitivity to Inhibitors of Nicotinamide Phosphoribosyltransferase in Acute Myeloid Leukemia"

### Supplemental Tables

Supplemental Table 1. Clinical characteristics of AML sample cohort

| | | Total (N = 270) | -7/-7q samples (n = 21; 7.8%) | WT samples (n = 249; 92.2%) | *p* |
|---|---|---|---|---|---|
| **Sex*** | Female, n (%) | 126 (46.7%) | 8 (38.1%) | 118 (47.4%) | |
| | Male, n (%) | 116 (43%) | 10 (47.6%) | 106 (42.6%) | NS |
| | NA, n (%) | 28 (10.4%) | 3 (14.3%) | 25 (10%) | |
| **Median age, years (range)†** | 29 (10.7%) NA | 62 (7–81) | 61.5 (22–71) | 62 (7–81) | NS |
| **AML Type*** | De novo, n (%) | 179 (66.3%) | 11 (52.4%) | 168 (67.5%) | |
| | sAML/tAML, n (%) | 84 (31.1%) | 9 (42.9%) | 75 (30.1%) | |
| | NA | 7 (2.6%) | 1 (4.8%) | 6 (2.4%) | NS |
| **Disease Stage*** | Diagnosis, n (%) | 129 (47.8%) | 8 (38.1%) | 121 (48.6%) | |
| | Relapse, n (%) | 91 (33.7%) | 7 (33.3%) | 84 (33.7%) | |
| | Refractory, n (%) | 44 (16.3%) | 6 (28.6%) | 38 (15.3%) | |
| | Remission, n (%) | 4 (1.5%) | 0 (0%) | 4 (1.6%) | NS |
| | NA, n (%) | 2 (0.7%) | 0 (0%) | 2 (0.8%) | |
| **Tissue Type*** | Bone marrow, n (%) | 242 (89.6%) | 16 (76.2%) | 226 (90.8%) | |
| | Blood, n (%) | 28 (10.4%) | 5 (23.8%) | 23 (9.2%) | NS |
| **Median blast % (range)†** | 27 (10%) NA's | 50 (3-100) | 66 (14–95) | 50 (3–100) | 0.041 |
| **FAB*** | AML M0, n (%) | 6 (2.2%) | 0 (0%) | 6 (2.4%) | |
| | AML M1, n (%) | 42 (15.6%) | 4 (19%) | 38 (15.3%) | |
| | AML M2, n (%) | 59 (21.9%) | 1 (4.8%) | 58 (23.3%) | |
| | AML M4, n (%) | 16 (5.9%) | 1 (4.8%) | 15 (6%) | |
| | AML M5, n (%) | 40 (14.8%) | 2 (9.5%) | 38 (15.3%) | |
| | AML M6, n (%) | 1 (0.4%) | 0 (0%) | 1 (0.4%) | |
| | AML M7, n (%) | 2 (0.7%) | 1 (4.8%) | 1 (0.4%) | NS |
| | NA, n (%) | 104 (38.5%) | 12 (57.1%) | 92 (36.9%) | |

FAB: French-American-British classification, sAML; secondary AML, tAML: therapy-related AML
NS: not significant, NA: data not available, WT: wildtype
* Fisher's exact test
† Mann–Whitney *U* test

Supplemental Table 2. Number of samples per experiment

| Experiment | All AML | -7/-7q | WT | Healthy |
|---|---|---|---|---|
| Total | 270 | 21 | 249 | 13 |
| Ex vivo drug sensitivity testing | 207 | 21 | 186 | 13 |
| Cell Titer-Glo | 189 | 21 | 168 | 6 |
| CellTox Green | 146 | 10 | 128 | 8 |
| Multi-parametric flow cytometry based drug sensitivity testin | 62 | 11 | 51 | 0 |
| Bulk RNA sequencing (FIMM cohort) | 183 | 12 | 171 | 0 |
| Single-cell RNA sequencing | 8 | 1 | 7 | 0 |

AML: acute myeloid leuekmia, WT: wildtype, FIMM: Institute for Molecular Medicine Finland

Supplemental Table 3. Flow cytometry antibody panel

| Antibody | Fluorophore | Clone | Isotype | Cell type specificity | Dilution factor | Vendor | Catalogue No. |
|---|---|---|---|---|---|---|---|
| CD45 | BV786 | HI30 | IgG1 Kappa | Hematopoietic cells | 1:100 | BD Horizon | 563716 |
| CD34 | APC | 8G12 | IgG1 Kappa | Myeloid progenitors | 1:100 | BD Biosciences | 345804 |
| CD38 | BV421 | HIT2 | IgG1 Kappa | Differentiated myeloid | 1:50 | BD Horizon | 562444 |
| CD14 | FITC | MϕP9 | IgG2b Kappa | Monocytes | 1:100 | BD Biosciences | 345784 |
| CD3 | PE-Cy7 | SK7 | IgG1 Kappa | Lymphocytes | 1:100 | BD Biosciences | 557851 |
| Annexin V | PE | - | - | Apoptotic cells | 1:50 | BD Pharmingen | 559763 |
| 7AAD | - | - | - | Dead cells | 1:50 | BD Pharmingen | 559763 |

### Supplemental Figures

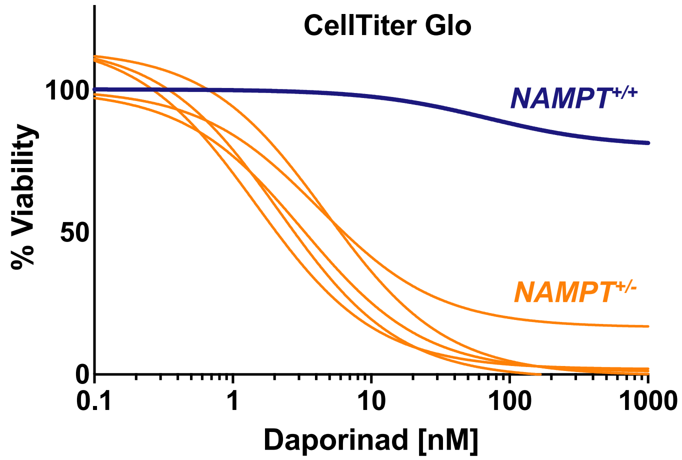


**Supplemental Figure 1**. AML with 7q deletion that does not encompass NAMPT lacks sensitivity to NAMPT inhibition. Ex-vivo daporinad dose response of an AML sample with -7q with a diploid *NAMPT* locus (*NAMPT*$^{+/+}$) compared with AML samples with *NAMPT* haploinsufficiency (*NAMPT*$^{+/-}$). Fresh bone marrow mononuclear cells were suspended in Mononuclear Cell Medium (PromoCell) and incubated with daporoinad for 72 h. Cell viability was measured by CellTiter-Glo.

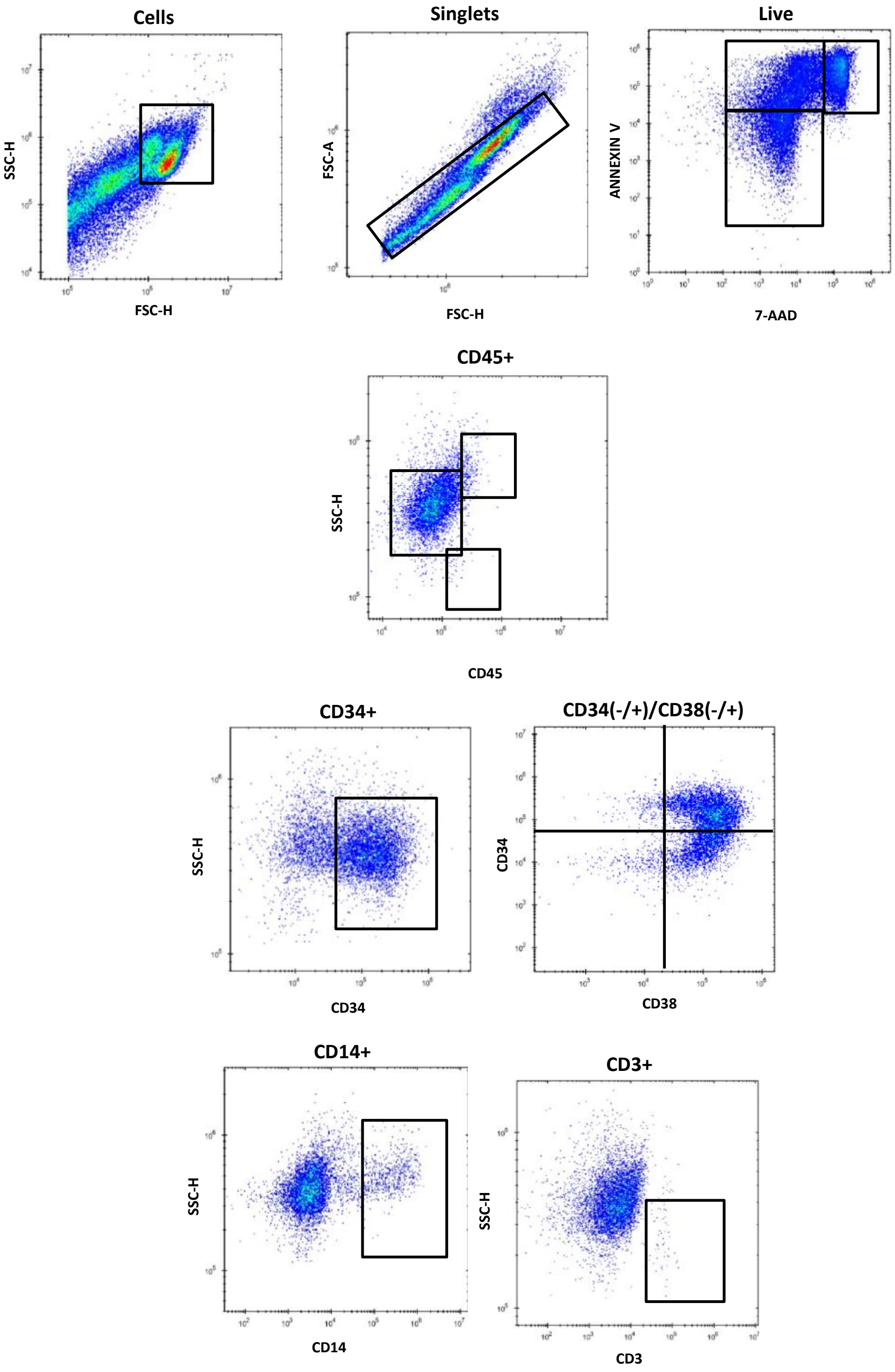


**Supplemental Figure 2.** Gating strategy used in drug sensitivity testing using multi-parametric flow cytometry.

### Supplemental methods

#### Patient samples

Bone marrow (BM) or peripheral blood (PB) samples were collected and mononuclear cells (MNCs) were isolated by Ficoll density gradient (Ficoll-Paque PREMIUM; GE Healthcare, Little Chalfont, Buckinghamshire, UK). MNCs were suspended in conditioned medium (CM) (RPMI 1640, 12,5% HS-5 conditioned medium, 10% FBS, 2 mM-glutamine, 100 units/ml penicillin, and 100 µg/ml streptomycin) [1]. An aliquot of MNCs were used for ex vivo drug sensitivity testing right after isolation and a portion of the cells was viably frozen.

#### Ex vivo drug sensitivity testing with patient cells

We evaluated the drug sensitivity of fresh MNCs to up to 527 small molecule inhibitors, including the NAMPT inhibitor daporinad. Cell viability and cytotoxicity were measured using the CellTiter-Glo (CTG) and CellTox Green (CTxG) (Promega) assays as previously described.[2–4] To test multiple different NAMPT inhibitors, daporinad (Axon Medchem), GMX1778 (Selleck Chemicals), KPT9274 (Selleck Chemicals), and LSN3154567 (Sigma-Aldrich) were dissolved in dimethylsulfoxide (DMSO) and dispensed on 384-well flat-bottom plates using the Echo 550 Acoustic Dispenser (Labcyte, San Jóse, CA, USA) in eight increasing concentrations. The concentration range tested for Daporinad, GMX1778, and LSN3154567 was 0.1-1000 nM, and for KPT-9274, 1-10000 nM. 0.1% DMSO and 100µM benzethonium chloride (BzCl) were used as negative and positive controls, respectively. Frozen MNCs from patients with AML were thawed and treated with RQ1 RNase-free DNase (1U/µl, Promega) to degrade DNA released from dead cells, and cells were left to recover for 4 h in 12.5% CM. Cells were plated onto pre-drugged plates in a concentration of 10000 cells/well. Cells were incubated with the drugs for 72 hours at 37°C and 5% $CO_2$. After the incubation, 25µl of CTG reagent (Promega) was added per well, and cells were incubated for 20 min at room temperature (RT). Luminescence was measured with a PHERAstar plate reader (BMG Labtech). Viability readouts per drug concentration were normalized to the mean of negative (DMSO) and positive (BzCl) controls. A four-parameter dose-response curve was fitted to the drug sensitivity data to define dose-response.

**Ex vivo drug sensitivity testing by multiparametric flow cytometry**

Daporinad, GMX1778, KPT9274, and LSN3154567 were pre-plated on Nunc™ 96-Well Polystyrene V-bottom plates (Thermo Fisher Scientific, Carlsbad, CA) using the Echo 550 Acoustic Dispenser in the same concentration range as previously. Thawed MNCs from AML patients were plated onto the pre-drugged plates in a concentration of 50,000 cells/well in 12.5% CM. After 72-hour incubation (37°C, 5% $CO_2$), cells were stained with antibodies against CD45, CD34, CD38, CD14, and CD3 (supplemental Table 3) (BD Biosciences, San Diego, CA, USA). To identify apoptotic and dead cells, the cells were stained with PE Annexin V and 7-amino actinomycin D (7-AAD) in Annexin V Binding Buffer (BD Biosciences). Cell viability was determined using the iQue Screener PLUS (Intellicyt, Albuquerque, NM, USA) flow cytometer. All cells were extracted from each well, and cell populations were gated using the ForeCyt software (Intellicyt). The gating strategy is shown in supplemental Figure 2. Drug sensitivity analysis was performed on the viable CD45-positive singlet cells by converting cell population counts to dose-response curves.

To determine the sensitivity of -7/-7q AML to combinations of the NAMPT inhibitor KPT-9274 and the BCL2 inhibitor venetoclax, we tested samples from three patients with AML who had an immature blast population (defined as CD34+CD38-) as determined by flow cytometry. Viably frozen bone marrow MNCs were thawed and incubated with either a single dose of KPT-9274 at 100nM, a single dose of venetoclax at 30nM, or both for 72 h at 37 °C. Afterward, the viability of the CD34+CD38- population was determined using the multiparametric flow cytometry assay.

**Quantification of drug sensitivity using Drug Sensitivity Score**

To facilitate statistical analysis of drug sensitivities, the dose-response curves were transformed into Drug Sensitivity Scores (DSS). For each dose-response curve, DSS was calculated using an integral function as previously described.[5] DSS is a metric that quantifies the area delineated by the dose-response curve, and that incorporates information on the compound's efficacy and potency. DSS values range from 0 to 50, 50 being the most sensitive.

**Drug sensitivity testing in cell lines**

AML cell lines were obtained from the German collection of microorganisms and cell cultures (DSMZ). The HL-60 and HL-60_TB cell lines were from the American type tissue culture collection (ATCC) and the NCI-Frederick cancer DCTD tumor/cell line repository, respectively. The KO52 cell line was from the JCRB Cell Bank (Osaka, Japan). AML cell lines were cultured in the vendor-recommended medium. Drug sensitivity testing was performed similarly to AML patient samples using CTG-based measurements. Daporinad was tested at five different concentrations (0.1 nM, 1 nM, 10 nM, 100 nM, and 1000 nM), and DMSO and BzCl were used as controls. 1500 to 3500 cells were plated per well using Multidrop Combi (Thermo Fisher Scientific) on pre-drugged 384-well flat-bottom plates.

**Identification of monosomy 7 and del(7q)**

Clinical karyotype data for AML patient samples were retrieved from the Finnish Hematology Registry and Biobank and from the Hospital District of Helsinki and Uusimaa hematological datalake. For AML samples without karyotype data, chromosomal copy number aberrations were identified based on exome sequencing of BM or PB and a matched skin biopsy sample. Whole exome libraries were prepared using NimbleGen SeqCap EZ v2.0, NimbleGen SeqCap EZ MedExome (Roche), Agilent SureSelect Target Enrichment System (Agilent), or Nextera Flex Exome (Illumina) kits according to the manufacturer's guidelines. The exome libraries were sequenced using a HiSeq 2500 instrument (Illumina, San Diego, CA, USA). Copy number aberrations were identified as described previously.[6] Copy number profiles for chromosome 7 were visually evaluated to identify -7 or del(7q).

Cell lines with -7/-7q were identified based on karyotype data provided by the vendor and by analysis of copy number aberration data from the Cancer Cell Line Encyclopedia.[7]

**RNA-sequencing and gene expression analysis**

TCGA gene expression values were retrieved from cBioportal[8] For the FIMM AML cohort, RNA extraction, library preparation, data preprocessing, and the analysis pipeline were performed as previously described.[9]

**Single-cell sequencing and data analysis**

To evaluate gene expression in bone marrow cell populations in patients with AML, we analyzed single-cell RNA-sequencing data obtained from viably frozen bone marrow mononuclear cells from eight patients. Sequencing and data processing for two samples (6333 and 5249) has been previously described.[10] The remaining six samples were sequenced and processed as follows. Samples were thawed in 12.5% CM, followed by washing and resuspension in 1X PBS with 0.04% BSA, before proceeding with 10x Genomics Single Cell Protocol. Single-cell gene expression profiles were determined using the 10X Genomics Chromium Single Cell 3'RNAseq platform (10x Genomics, CA, USA). The gel beads in emulsion (GEM) generation, cDNA amplification, and library preparation were performed using the Chromium Next GEM Single Cell 3' Gene Expression version 3.1 Dual Index Reagent kit (10x Genomics). The Sample libraries were sequenced on Illumina NovaSeq 6000 system using read lengths 28 bp (Read 1), 10 bp (i7 Index), 10 bp (i5 Index), and 90 bp (Read 2). The Cell Ranger mfastq and count pipelines (version 6.0.0.) were used to generate FASTQ files and to perform alignment, filtering, and UMI counting, respectively. Reads were aligned to reference genome GRCh38. Single-cell data was analyzed using Seurat v4.3.0[11] and R 4.2.1. Cells with > 20% mitochondrial gene counts, or less than 200 detected genes or low gene complexity (log10GenesPerUMI <0.8) were filtered out to remove cells with poor quality data. Samples were integrated using the reciprocal PCA method to correct for technical differences between datasets and to perform comparative single-cell RNA sequence analysis between different samples. Cell types were identified using the ScType pipeline, supplemented with in-house gene sets.[12]

### Statistical Tests

Statistical analyses were performed in the R Software, and nonparametric tests were used when values did not follow a normal distribution. Comparisons of DSS scores between two sample groups were made with either the 2-sample t-test or the Mann-Whitney U-test.

### References


1. Karjalainen R, Pemovska T, Popa M, et al. JAK1/2 and BCL2 inhibitors synergize to counteract bone marrow stromal cell–induced protection of AML. *Blood*. 2017;130(6):789–802.
2. Malani D, Kumar A, Brück O, et al. Implementing a Functional Precision Medicine Tumor Board for Acute Myeloid Leukemia. *Cancer Discovery*. 2022;12(2):388–401.
3. Pemovska T, Kontro M, Yadav B, et al. Individualized Systems Medicine Strategy to Tailor Treatments for Patients with Chemorefractory Acute Myeloid Leukemia. *Cancer Discov*. 2013;3(12):1416–1429.
4. Tambe M, Karjalainen E, Vähä-Koskela M, et al. Pan-RAF inhibition induces apoptosis in acute myeloid leukemia cells and synergizes with BCL2 inhibition. *Leukemia*. 2020;34(12):3186–3196.
5. Yadav B, Pemovska T, Szwajda A, et al. Quantitative scoring of differential drug sensitivity for individually optimized anticancer therapies. *Sci Rep*. 2014;4:5193.
6. Eldfors S, Kuusanmäki H, Kontro M, et al. Idelalisib sensitivity and mechanisms of disease progression in relapsed TCF3-PBX1 acute lymphoblastic leukemia. *Leukemia*. 2017;31(1):51–57.
7. Barretina J, Caponigro G, Stransky N, et al. The Cancer Cell Line Encyclopedia enables predictive modelling of anticancer drug sensitivity. *Nature*. 2012;483(7391):603–607.
8. Gao J, Aksoy BA, Dogrusoz U, et al. Integrative Analysis of Complex Cancer Genomics and Clinical Profiles Using the cBioPortal. *Science Signaling*. 2013;6(269):pl1–pl1.
9. Kumar A, Kankainen M, Parsons A, et al. The impact of RNA sequence library construction protocols on transcriptomic profiling of leukemia. *BMC Genomics*. 2017;18(1):629.
10. Dufva O, Pölönen P, Brück O, et al. Immunogenomic Landscape of Hematological Malignancies. *Cancer Cell*. 2020;38(3):380-399.e13.
11. Hao Y, Hao S, Andersen-Nissen E, et al. Integrated analysis of multimodal single-cell data. *Cell*. 2021;184(13):3573-3587.e29.
12. Ianevski A, Giri AK, Aittokallio T. Fully-automated and ultra-fast cell-type identification using specific marker combinations from single-cell transcriptomic data. *Nat Commun*. 2022;13(1):1246.
